# Menstrual Cycle Phase-Based Interval Training Yields Distinct Muscle Changes in Female Athletes

**DOI:** 10.1101/2024.10.28.24316287

**Authors:** Julie Kissow, Kamine J. Jacobsen, Søren Jessen, Laura B. Thomsen, Júlia P. Quesada, Jens Bangsbo, Atul S. Deshmukh, Morten Hostrup

**Affiliations:** The August Krogh Section for Human Physiology, Department of Nutrition, Exercise and Sports, University of Copenhagen, Denmark; The Novo Nordisk Foundation Center for Basic Metabolic Research, Faculty of Health and Medical Sciences, University of Copenhagen, Copenhagen, Denmark

**Keywords:** Estrogen, Exercise, Female, Performance, Sex hormones

## Abstract

Advances in mass-spectrometry(MS)-based technologies have leveraged our understanding of skeletal muscle responsiveness to exercise in humans. However, there is a lack of such data in females, particularly pertaining to female athletes and menstrual cycle phase-based sprint interval training(SIT) despite its efficacy and popularity. Here, we present a comprehensive proteomic analysis of skeletal muscle adaptations to high-frequency SIT during different menstrual cycle phases in female athletes. We randomized 49 eumenorrheic females to either high-frequency SIT in the follicular(FB) or luteal phase(LB) over one menstrual cycle comprising eight sessions of 6×30-s all-out efforts. Twenty-five completed the intervention with verified cycles. MS-based proteomics revealed notable differences in muscle adaptations to phase-based SIT. LB suppressed mitochondrial pathways of the tricarboxylic acid cycle and electron transport chain while enriching ribosomal complexes. Conversely, FB enriched filament organization and skeletal system development. Mitochondrial repression during LB was linked to reduced V̇O_2max_, whereas exercise capacity improved in FB only. Our findings show that synching high-frequency SIT with menstrual cycle phases induces distinct muscle adaptations and affects phenotype in eumenorrheic female athletes.

## Introduction

Skeletal muscle has a critical role in driving the health- and performance-related benefits of exercise (Pedersen & Febbraio, 2012). It exhibits a remarkable ability to adapt in response to even short periods of physical activity (Almquist et al., 2021; Iaia et al., 2008). Recent advances in mass-spectrometry (MS)-based technologies have leveraged the muscle physiological field during the past years by allowing in-depth coverage of thousands of proteins. Utilizing MS-based workflows, different research groups have revealed that only a few weeks of exercise training regulates several hundreds of proteins in human skeletal muscle (Aebersold & Mann, 2016; Deshmukh et al., 2021; Granata et al., 2021; Hostrup et al., 2022). However, a major limitation of these studies is the exclusion of female participants, nor were they performed in a trained cohort, thus neglecting physiological nuances (Ansdell et al., 2020). Investigating global protein-level adaptations in female participants would provide valuable insights and could reveal distinct regulation patterns in response to exercise training in females.

Exercise training studies in female athletes are massively underrepresented within the fields of sports medicine and exercise physiology (Cowley et al., 2021; Elliott-Sale et al., 2021). The common arguments for excluding females in exercise interventions often pertain to fluctuations in female hormones during the menstrual cycle potentially interfering with study outcomes. The impact of the menstrual cycle phases on female performance and training responsiveness has attracted increasing interest but revealed somewhat divided opinions (D’Souza et al., 2023; Janse et al., 2019; Kissow et al., 2022; McNulty et al., 2020; Thompson et al., 2020). While some categorically claim that the menstrual cycle has no impact, a few studies indicate that phase-based training – that is, training primarily in the follicular or luteal phase of the menstrual cycle – may affect the response to resistance training (Gunnarsson & Bangsbo, 2012; Reis et al., 1995; Sakamaki-Sunaga et al., 2016; Sung et al., 2014; Wikstrom-Frisen et al., 2017). However, knowledge about intrinsic training adaptations within skeletal muscle is absent and studies on the impact of phase-based interval training are lacking despite being an integral part of weekly training programs. This is particularly true for sprint interval training (SIT), which is an increasingly popular form of interval training performed at very high intensities (Hazell et al., 2010; Hostrup et al., 2021; Yamagishi & Babraj, 2017). In spite of multiple studies showing beneficial performance adaptations to SIT in already well-trained individuals (Hostrup & Bangsbo, 2017), only a few studies have been conducted with trained females (Cicioni-Kolsky et al., 2013; Vandbakk et al., 2017), and none of these assessed the influence of the menstrual cycle on training outcomes (Ikeda et al., 2019; Janse et al., 2019).

The rationale for the menstrual cycle to affect training outcomes resides in hormonal fluctuations. In the early-follicular phase, estrogen and progesterone levels are low, contrasting the late-follicular phase marked by increased estrogen release triggered by follicle-stimulating hormone (FSH) and luteinizing hormone (LH) secretion, leading to ovulation. Subsequently, in the luteal phase both hormones are secreted (Oosthuyse & Bosch, 2010). Some evidence suggests that estrogen has an anabolic effect on skeletal muscle and thus can lead to an increased potential for training adaptations (Elliott-Sale et al., 2021; Hansen, 2018; Ikeda et al., 2019), possibly through estrogen-induced satellite cell activation post-exercise (Enns & Tiidus, 2008; Thomas et al., 2010). In accordance, a few studies found superior resistance training adaptations to follicular phase-based training than luteal phase-based training in terms of enhancing muscle strength and mass (Kissow et al., 2022; Thompson et al., 2020). In addition, artificial clamping of hormone levels with hormonal prevention methods are known to affect training outcomes (Dalgaard et al., 2019; Oxfeldt et al., 2020; Schaumberg et al., 2017). Therefore, it is conceivable that naturally occurring fluctuations in hormones during the menstrual cycle may affect training outcomes too.

The major challenge associated with investigating this topic is detailed tracking of individual participants’ menstrual cycle lengths and transitions from the follicular to luteal phase (Elliott-Sale et al., 2021; Janse et al., 2019). Calendar-based counting and basal body temperature to estimate day for ovulation are each associated with their own drawbacks and inaccuracies, mostly pertaining to inter- and even intra-individual differences in phase length. The most feasible and accurate solution to this challenge is a combination of urinary LH measurement monitoring, calendar-based counting and blood sampling for estrogen and progesterone (Janse et al., 2019). Yet, such a comprehensive phase assessment has never been undertaken, as most of the few available studies to date utilized only a single method for phase determination, thus leaving the important question of menstrual cycle phase and its effects on training outcomes inadequately explored.

Here, we employ a comprehensive MS-based proteomics workflow and strict set of inclusion criteria with longitudinal monitoring in a cohort of female athletes to uncover protein-wide muscle adaptations in response to high-frequency SIT in either the follicular or luteal phase of the menstrual cycle. Utilizing our workflow, we show that luteal phase-based SIT consistently downregulated mitochondrial proteins and proteins comprising the tricarboxylic acid cycle. In contrast, follicular phase-based SIT shows greater adaptation in extracellular matrix organization proteins and maintenance of all major metabolic pathways. Furthermore, we demonstrate superior follicular phase-based training effects in cardiorespiratory fitness, but not time trial performance. Collectively, these results suggest that menstrual cycle phase is an important confounder of muscle adaptations to exercise training.

## Results

### Participants exhibited regular menstrual cycles and trained in the correct phases

To investigate the influence of menstrual phase in the response to exercise training, we included female athletes and randomized them, stratified for V̇O_2max_, to high-frequency training in either their luteal (LB) or follicular phase (FB) according to a schedule personalized to the individual’s menstrual cycle. To verify the menstrual phases before and throughout the study, we utilized a comprehensive inclusion process (**Fig. 1**A). Before commencement of the first experimental trials, participants kept a 3-month detailed calendar-based menstrual cycle log to register onset of menses to verify regular length of the menstrual cycle (defined as 21-35 days). We also assessed urinary luteinizing hormone (LH) from day 9 of the last cycle before commencement in the study using daily ovulation kits to confirm the transmission from the follicular to the luteal phase (**Fig. 1**B). Furthermore, as proposed by Janse et al. (Janse et al., 2019), we quantified serum progesterone concentrations during experimental days and excluded participants with values below 16 nmol·L^-1^ from data analysis as this can indicate luteal phase deficiency. Utilizing these strict set of criteria, we initially screened 82 females of which 49 fulfilled the eligibility criteria for inclusion (**Fig. 1**C). Of these, 33 completed the training intervention of which eight was excluded due to serum progesterone concentrations <16 nmol·L^-1^ (FB n=4, LB n=4; **Fig. 1**D). Thus, the final analysis is based on the remaining 25 females (FB n=13, LB n=12) (Table 1) with confirmed cycle lengths of follicular and luteal phases of equal duration (**Fig. 1**E). Although cycle length varies (Dam et al., 2022), our included females exhibited little variation in phase length (follicular phase: 1±1 (mean ± SD) day; luteal phase: 1±1 day) and day of ovulation (1±1 day).

**Fig. 1.**
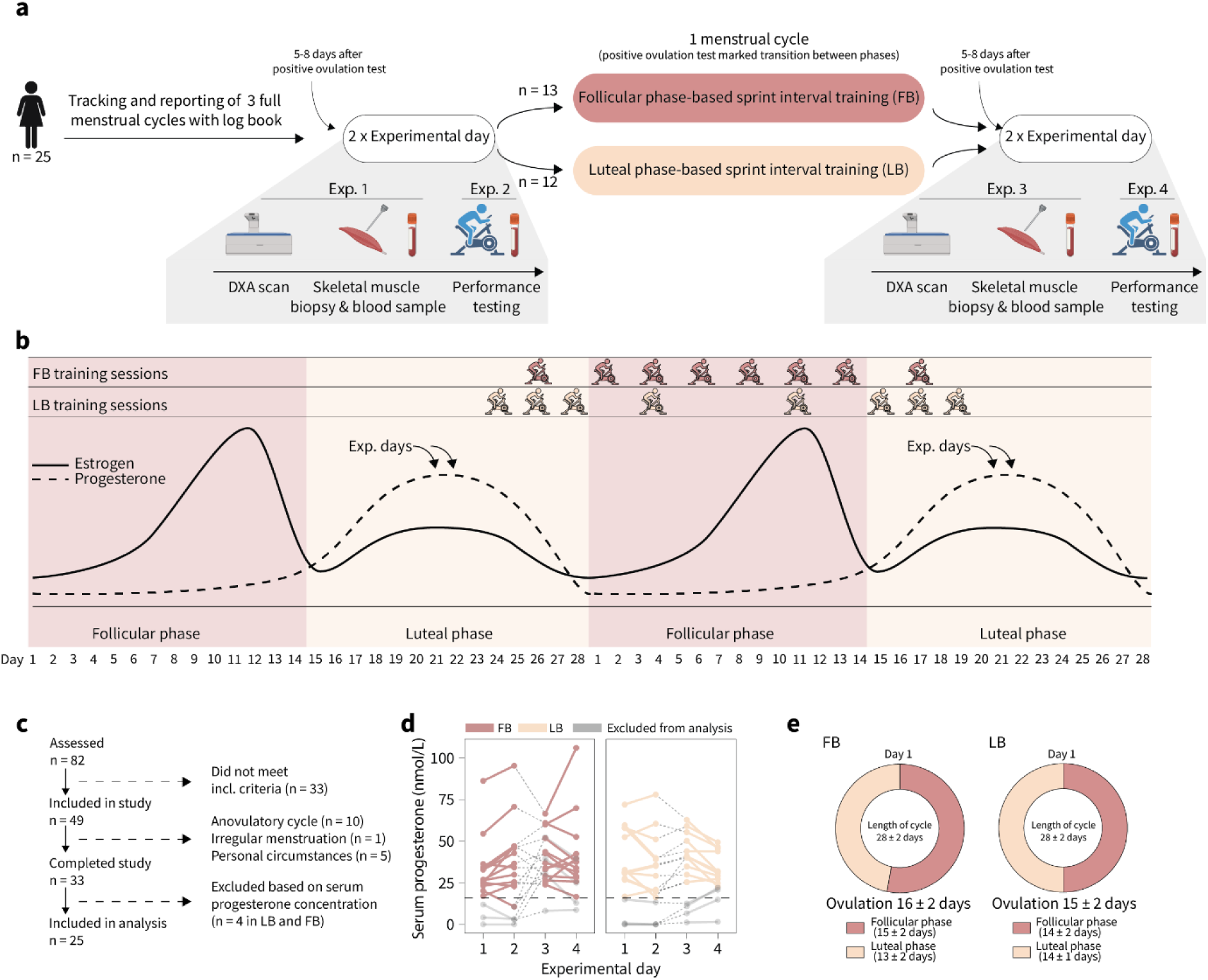
Experimental overview of high-frequency phase-based sprint interval training (SIT) in the follicular phase (FB) and luteal phase (LB). (**A**) Experimental overview of the study design. Participants tracked their menstrual cycle for 3 cycles prior to enrollment to ensure a regular menstrual cycle (between 21 and 35 days). (**B**) Overview of the menstrual cycle phase-based sprint interval training intervention. (**C**) Flow diagram of the inclusion process. (**D**) Serum progesterone concentrations on trial days. The lower limit on 16 nmol·L^-1^ is indicated by a dashed line, and participants who were below this limit on any one occasion were excluded from data analysis (marked by grey lines and symbols). Participants are marked by dashed lines between symbols between trial 2 and 3. (**E**) Mean length for the menstrual cycle in the two groups. DXA: Dual Energy X-Ray Absorptiometry; FB: follicular phase-based training; LB: luteal phase-based training.

**Table 1:** Characteristics of the participants in the follicular phase-based training (FB) and luteal phase-based training (LB).

|  | **FB** (*n*=13) | **LB** (*n*=12) | **ALL** (*n*=25) |
| --- | --- | --- | --- |
| Age (*years*) | 28 ± 3 | 29 ± 5 | 28 ± 4 |
| Weight (*kg*) | 60.3 ± 6.1 | 69.9 ± 7.7 | 64.9 ± 8.4 |
| BMI | 22.1 ± 1.0 | 24.5 ± 2.1 | 23.3 ± 2.0 |
| Fat percent (%) | 22.8 ± 3.2 | 23.8 ± 4.2 | 23.2 ± 3.8 |
| VO<sub>2max</sub> (*mL·min<sup>-1</sup>*) | 3107 ± 328 | 3485 ± 472 | 3288 ± 439 |
| VO<sub>2max</sub> (*mL·min<sup>-1</sup>·kg<sup>-1</sup>*) | 51.7 ± 3.5 | 49.9 ± 4.1 | 50.8 ± 3.9 |
| Training level (*hours·week<sup>-1</sup>*) | 5.9 ± 2.8 | 7.2 ± 4.1 | 6.5 ± 3.5 |
| BMI: body mass index; VO<sub>2max</sub>: maximal oxygen consumption. Data are presented as mean ± SD. |  |  |  |

During the 4-week SIT intervention, we determined menstrual cycle phases continuously by both calendar-based counting and ovulation test with the onset of testing from day 9 in the cycle until a positive test occurred. We did this to ensure the transition between the follicular and luteal phase during the intervention. The training intervention consisted of 8 SIT sessions on a spinning bike, each consisting of 6×30-s all out bouts interspersed by 3 min of rest. FB performed high-frequency SIT in the follicular phase (6 training sessions) and low-frequency SIT in the luteal phase (2 training sessions). In contrast, LB performed low-frequency SIT training in the follicular phase (2 training sessions) and high-frequency SIT in the luteal phase. (6 training sessions) (**Fig. 1**B). Experimental trials pre and post intervention were placed 5-8 days after positive ovulation (mid-luteal phase), where both progesterone and estrogen levels are expected to be at their highest and most stable (Oosthuyse & Bosch, 2010). We adopted this approach to more confidently attribute any differences between FB and LB to muscular adaptations from the training intervention, rather than to fluctuations in hormone concentrations on the specific experimental day (McNulty et al., 2020). This corresponded to the participants performing their post-experimental trials 48-72 hours after the last training session, hence allowing for sufficient recovery before experimental trials.

### Superior outcomes in cardiorespiratory fitness with follicular phase-based high-frequency SIT

Menstrual cycle phase-based SIT led to differential outcomes in cardiorespiratory fitness as reflected by between-group changes in V̇O_2max_ both in absolute terms and relative to body mass (group×time interaction: P=0.027) (**Fig. 1**A-B). This was related to a decline in V̇O_2max_ by 117 mL·min^-1^ in LB (95%CI −204 to −29, P=0.047), corresponding to −2.7%, whereas V̇O_2max_ was maintained in FB (P=0.364). While this was not as clearly reflected in terms of exercise capacity during the ramp test, only FB improved by 9 W (95% 0.3 to 18, P=0.043), whereas LB was unchanged (P=0.857; **Fig. 1**C). During a 30-min time trial, LB performed non-significantly greater after the intervention (P=0.076; **Fig. 2**G), which was generally related to a greater power output in the initial part of the test (**Fig. 1**F), whereas no apparent change was observed in FB (P=0.614). Body mass distribution did not change with both interventions (**Fig. 1**D-E).

**Fig. 2.**
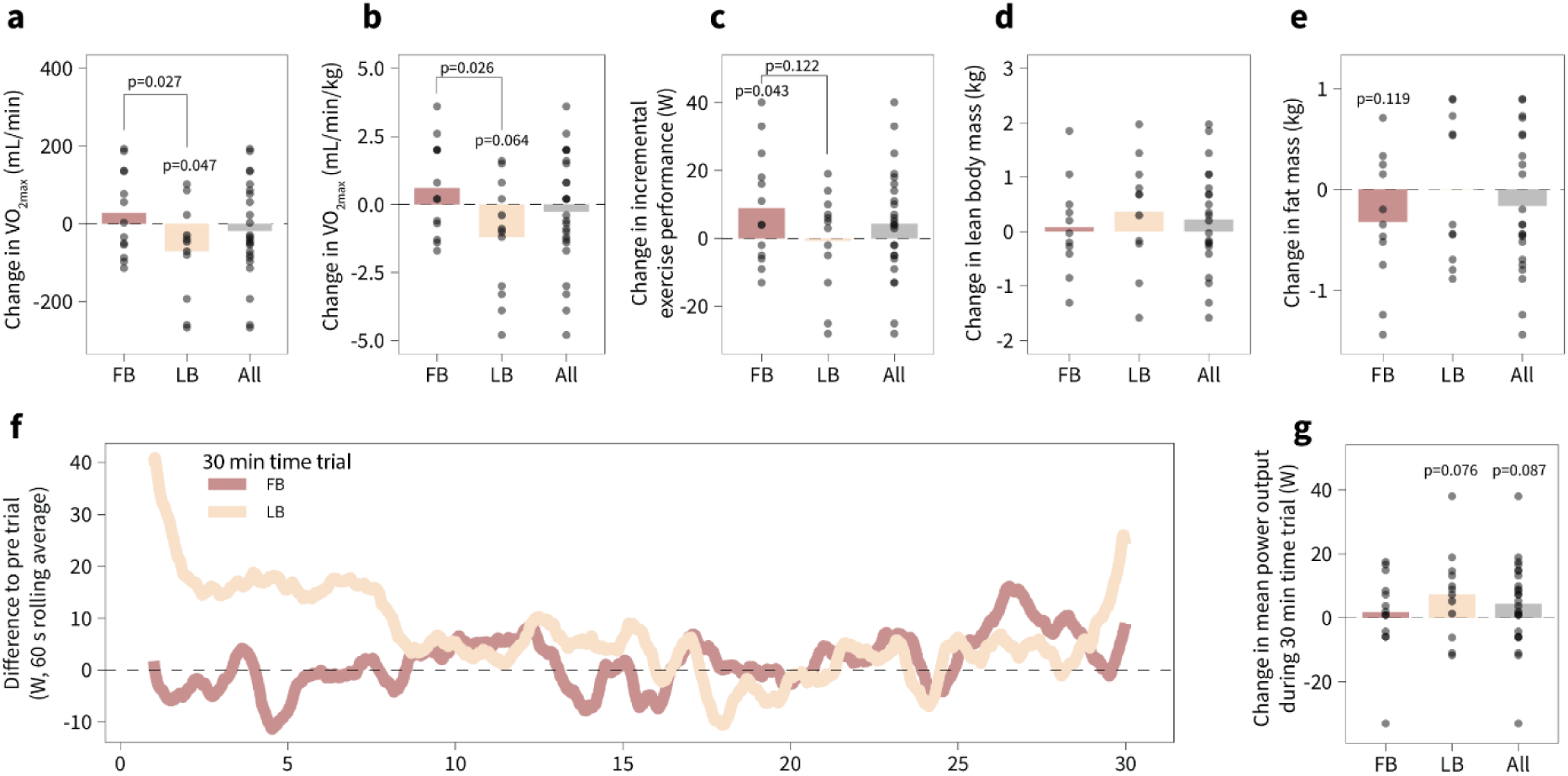
High-frequency phase-based sprint interval training (SIT) in the follicular phase (FB) and luteal phase (LB) induces differential outcomes in maximal oxygen consumption and incremental exercise performance. (**A**) Change in maximal oxygen uptake (V̇O_2max_). (**B**) Change in relative V̇O_2max_. (**C**) Change in incremental exercise performance. (**D**) Change in lean body mass. (**E**) Change in fat mass. (**F**) Mean power output during a 30-min all-out time trial. Lines are 60s rolling averages. (**G**) Change in mean power output during the 30-min time trial. Data are presented as means with individual changes. P-values above bars are within-group effect. P-values between treatments are between-group (group × time interaction effect). FB: follicular phase-based training; LB: luteal phase-based training.

### Menstrual cycle phase-based SIT induces differential muscle proteomic remodeling

To comprehensively decipher protein-wide adaptations in skeletal muscle in response to phase-based SIT, we utilized a state-of-the-art proteomics workflow (**Fig. 3**A). We identified ∼4300 proteins per sample (Table S2, **Fig. 3**B, 4155 after filtering for 70% valid values) with a dynamic range spanning 5 orders of magnitude (**Fig. 3**C). To assess within- and between-group changes, we employed a statistical approach combining biological relevance (log_2_fold change) as well as statistical significance (p-value) into a Π-score, as previously described (Peronnet & Massicotte, 1991), with a selected cut-off of 0.05 (**Fig. 3**D). In doing so, we uncovered a modest regulation in FB, with 10 proteins being differentially regulated, compared to 41 in LB (Table S2). Notably, the overlap in regulation was very small (**Fig. 3**E), and even when comparing the changes of all proteins irrespective of Π-score, correlation of changes between groups was low (**Fig. 3**F), demonstrating a notable divergence in muscle adaptation with phase-based training.

**Fig. 3.**
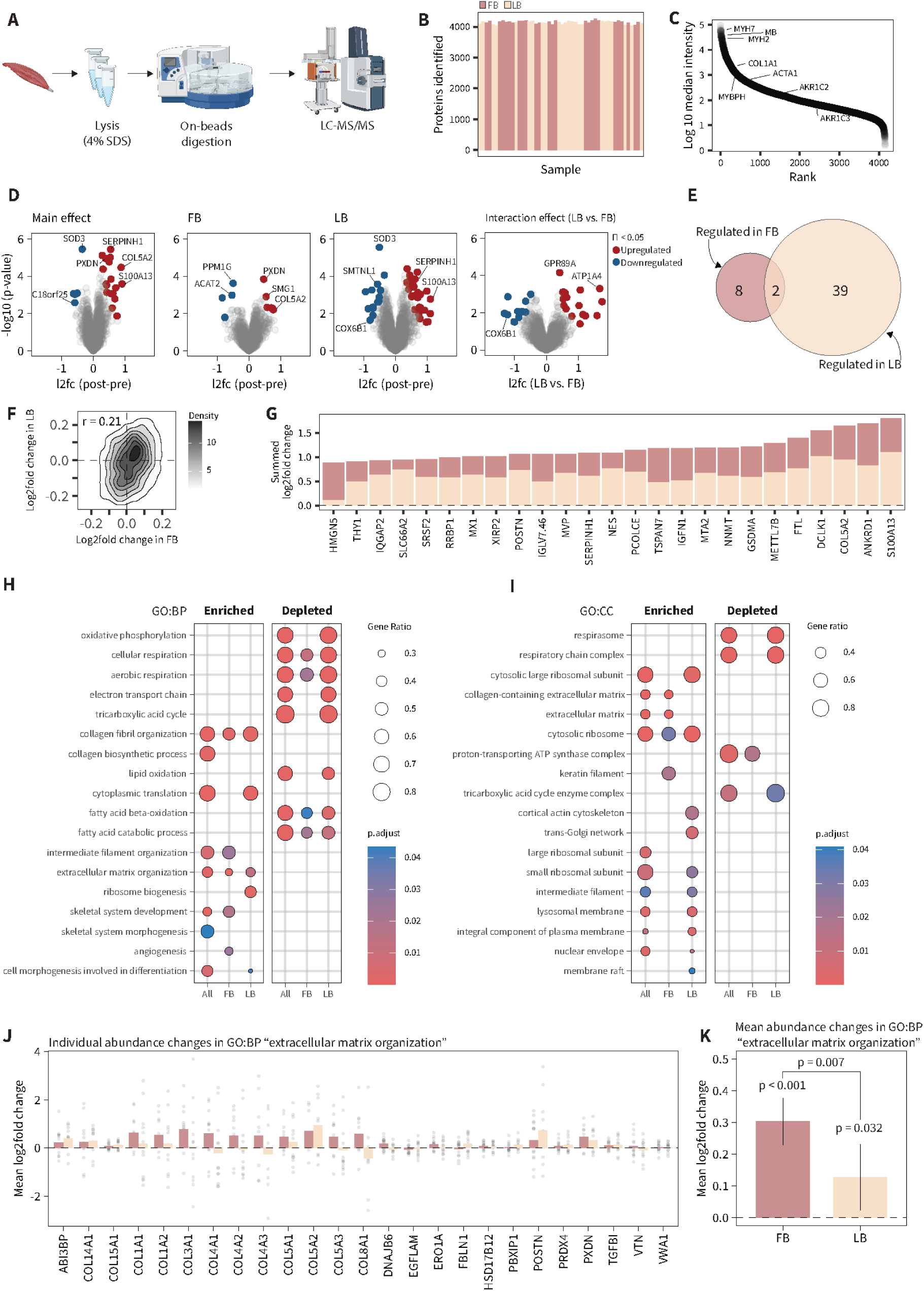
Four weeks high-frequency phase-based SIT does not maintain capacity for aerobic energy production and metabolic pathways in the luteal phase. (**A**) Overview of sample preparation. (**B**) Protein groups identified per sample. (**C**) Dynamic range of protein identifications. (**D**) Volcano plots of post-pre differences for the main effect, within-group effects, and interaction effects (LB – FB). (**E**) Venn diagram showing overlap in regulation between FB and LB. (**F**) Correlation density plot between proteomic changes in FB and LB with Pearson correlation coefficient. (**G**) Summed mean log2fold changes with the training interventions. (**H**) Gene set enrichment analysis of biological processes. (**I**) Gene set enrichment analysis of cellular compartments. Panels H and I both analyzed with clusterProfiler (v. 4.6.2). (**J**) Individual log2fold changes of extracellular matrix organization proteins. (**K**) mean log2fold changes of extracellular matrix organization proteins.

Among the twenty-five most upregulated candidates were proteins such as SERPINH1 and PCOLCE (both involved in the biosynthetic pathway of collagen (Casazza et al., 2002; Dean et al., 2003)), COL5A2 (structural component of group 1 collagen), FTL (involved in iron homeostasis), and S100A13 (a member of the calcium-binding S100 super family)(**Fig. 3**G) suggesting adaptations in tissue remodeling and cellular signaling pathways across both phase-based training interventions.

To further explore the biological processes affected by phase-based training, we performed gene set enrichment analysis. Surprisingly, the most significant changes were terms related to mitochondrial energy production, in which luteal phase-based training appeared to exhibit a pronounced depletion, which was paralleled by depletion in proteins related to the tricarboxylic acid cycle (**Fig. 3**H-I). On the other hand, both groups exhibited enrichment in terms related to ribosomal biogenesis, likely reflecting an increased need for protein turnover in response to the training intervention. In addition, extracellular matrix organization, particularly protein of the major collagen types, were upregulated in both groups (**Fig. 3**J) with a greater increase with follicular phase-based training (**Fig. 3**K).

### Luteal phase-based training does not maintain mitochondrial protein abundance or proteins of the tricarboxylic cycle

To unfold the gene set enrichment analysis findings, we assessed all proteins belonging to a mitochondrial complex of the electron transport chain (annotated with MitoCarta 3.0). Here, we were surprised to note a consistent decrease in abundance of proteins belonging to complex I-IV of the electron transport chain with luteal phase-based training (**Fig. 4**A), suggesting impaired mitochondrial energy production and metabolic efficiency, which coincides with the observed decrease in V̇O_2max_. Distinct responses between follicular and luteal phase-based training were also evident for proteins of the tricarboxylic acid cycle, and to a lesser extent fatty acid oxidation (**Fig. 4**B), highlighting the distinct metabolic adaptation to phase-based training. However, proteins of the glycolysis pathway were seemingly unaffected by group. And while mitochondrial and TCA pathways were detrimentally affected in the LB group, cytosolic ribosomal proteins exhibited increases in abundance which was favorable towards luteal phase-based training (**Fig. 4**C).

**Fig. 4.**
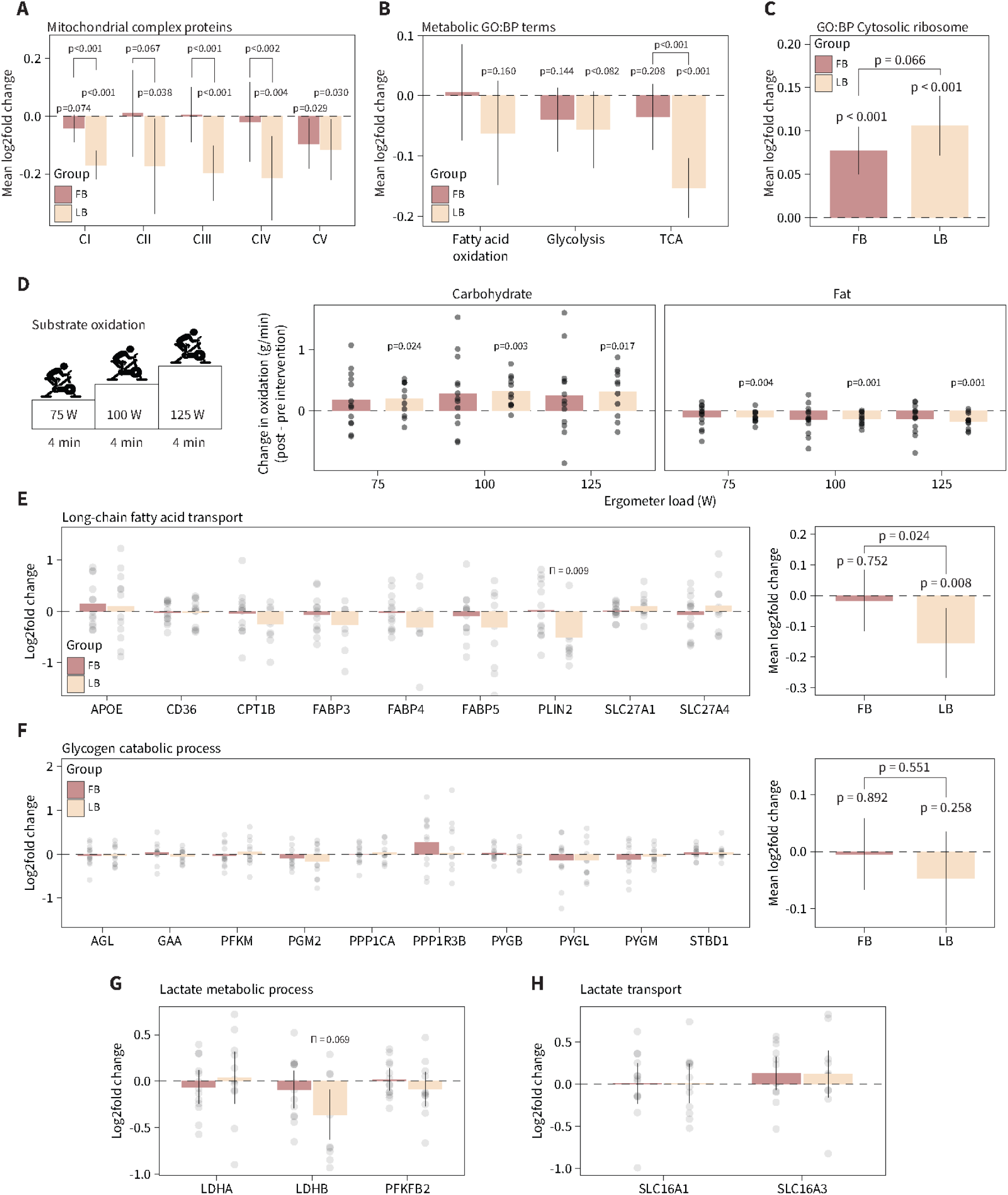
(**A**) Mean log2fold changes in mitochondrial protein complex abundances. Proteins were annotated with MitoCarta 3.0. (**B**) Mean log2fold changes in abundances of proteins annotated to gene ontology terms related to fatty acid oxidation, glycolysis, and tricarboxylic acid cycle. (**C**) Mean log2fold changes in abundances of proteins annotated to gene ontology term “cytosolic ribosome”. (**D**) Substrate oxidation (carbohydrate, left panel; fat, right panel) determined by indirect calorimetry. (**E**) Individual (left panel) and mean (right panel) log2fold changes of proteins annotated to go:bp term “long-chain fatty acid transport”. (**F**) Individual (left panel) and mean (right panel) log2fold changes of proteins annotated to go:bp term “glycogen catabolic process”. (**G**) Individual log2fold changes for go:bp term “lactate metabolic process”. (**H**) Individual log2fold changes for go:bp term “lactate transport”. P-values above bars are within-group and between treatments are between-group (group × time interaction effect). FB: follicular phase-based training; LB: luteal phase-based training.

We investigated training induced changes in substrate oxidation, which was not affected by phase-based training (**Fig. 4**D). However, we observed an overall training effect across all the three submaximal loads, with an increase in carbohydrate oxidation (P = 0.024) and a concomitant decrease in fat oxidation (P≤0.001, Table 2). At all three submaximal loads in LB, carbohydrate oxidation was increased and fat oxidation decreased (**Fig. 4**D). In comparison, there were no changes in substrate oxidation with FB. Intriguingly, this increased reliance on carbohydrate metabolism in LB coincides with overall reduction in mitochondrial proteins with decreased ability to oxidize fat as consequence (Spriet, 2014), as well as overall greater reductions in proteins related to long-chain fatty acid transport in LB (**Fig. 4**E). Interestingly, pathways of importance for short-term high-intensity energy production, such as glycogenolysis (**Fig. 4**F) and lactate metabolism and transport (**Fig. 4**G-H), were not affected by SIT or menstrual phase-based. The change in oxidation rates in LB display accurate biological coherence with impaired mitochondrial and metabolic pathway protein abundances, as well as V̇O_2max_ and incremental exercise performance, demonstrating differential skeletal muscle training adaptation incurred by menstrual cycle phase-based training.

**Table 2.** Substrate oxidation during submaximal exercise after 4 weeks of sprint interval training with either follicular phase-based training (FB) or luteal phase-based training (LB).

|  | Carbohydrate oxidation (g/min) |  |  | Fat oxidation (g/min) |  |  |
| --- | --- | --- | --- | --- | --- | --- |
|  | 75 (W) | 100 (W) | 125 (W) | 75 (W) | 100 (W) | 125 (W) |
| **FB (n=13)** |  |  |  |  |  |  |
| Pre | 1.16 ± 0.29 | 1.61 ± 0.32 | 2.11 ± 0.41 | 0.33 ± 0.16 | 0.30 ± 0.17 | 0.25 ± 0.18 |
| Post | 1.34 ± 0.36 | 1.90 ± 0.45 | 2.36 ± 0.48 | 0.24 ± 0.13 | 0.16 ± 0.16 | 0.12 ± 0.15 |
| $\Delta$ (95%CI) | 0.18 (-0.08 to 0.44) | 0.29 (-0.05 to 0.63) | 0.26 (-0.12 to 0.63) | -0.09 (-0.20 to 0.02) | -0.13 (-0.27 to 0.00) | -0.13 (-0.26 to 0.01) |
| **LB (n=12)** |  |  |  |  |  |  |
| Pre | 0.98 ± 0.31 | 1.50 ± 0.21 | 1.87 ± 0.34 | 0.43 ± 0.15 | 0.34 ± 0.11 | 0.35 ± 0.15 |
| Post | 1.21 ± 0.34 | 1.80 ± 0.30 | 2.18 ± 0.40 | 0.31 ± 0.13 | 0.20 ± 0.13 | 0.18 ± 0.16 |
| $\Delta$ (95%CI) | 0.21 (0.03 to 0.39)* | 0.31 (0.15 to 0.48)* | 0.31 (0.07 to 0.56)* | -0.10 (-0.16 to -0.04)* | -0.13 (0.13 to -0.06)* | -0.17 (-0.25 to -0.09)* |
| **ALL (n=25)** |  |  |  |  |  |  |
| Pre | 1.07 ± 0.31 | 1.55 ± 0.27 | 2.00 ± 0.39 | 0.38 ± 0.16 | 0.32 ± 0.15 | 0.29 ± 0.17 |
| Post | 1.28 ± 0.35 | 1.85 ± 0.38 | 2.28 ± 0.44 | 0.27 ± 0.13 | 0.18 ± 0.15 | 0.15 ± 0.15 |
| $\Delta$ (95%CI) | 0.19 (0.03 to 0.35)* | 0.30 (0.11 to 0.50)* | 0.28 (0.06 to 0.51)* | -0.09 (-0.16 to -0.03)* | -0.13 (-0.21 to -0.06)* | -0.14 (-0.23 to -0.06)* |

## Discussion

Herein, we utilized MS-based proteomics and a strict set of inclusion criteria with longitudinal monitoring in a cohort of eumenorrheic female athletes to uncover protein wide adaptations in skeletal muscle in response to high-frequency follicular- or luteal phase-based SIT. While SIT, in general, enriched ribosomal content, we observed some notable divergent adaptations between the phase-based training groups. Specifically, we show that luteal phase-based SIT elicits pronounced dilution of the mitochondrial pool and downregulation of tricarboxylic acid cycle proteins, and on the whole-body level, a decrease in cardiorespiratory fitness. In contrast, follicular phase-based training elicited superior adaptations in extracellular matrix reorganization and maintenance of the major metabolic pathways. These findings provide intriguing indications that exercise responsiveness may be heightened during the high frequency training sessions performed in the follicular phase of the menstrual cycle in female athletes.

SIT is a widely utilized training form because of its demonstrated efficacy to enhance performance in the severe intensity domain in already well-trained individuals and its efficiency to promote beneficial cardiovascular and mitochondrial adaptations within only a few weeks in sedentary individuals (Edge et al., 2013; Girard et al., 2011; Hostrup & Bangsbo, 2017). Here, we provide the first in-depth proteome analysis of the protein-wide response to SIT in skeletal muscle of a female athlete cohort. Despite the already trained nature of the included cohort, we show that SIT triggers pronounced upregulation of ribosomal and extracellular matrix proteins, likely in response to increased demands for protein synthesis and structural integrity (Brightwell et al., 2022; Kritikaki et al., 2021). Delving deeper, we reveal pronounced differences in energy production pathways depending on high-frequency training distributions to the follicular or luteal phase. This was particularly evident for proteins of the oxidative phosphorylation complexes, which exhibited a clearly pronounced decrease with luteal phase-based training for all complexes (CI-CV). This was paralleled by a consistent suppression of proteins related to the tricarboxylic acid and lipid metabolism of which PLIN2 was the most repressed protein. In contrast, follicular phase-based training maintained oxidative phosphorylation complex abundance and metabolic pathways, demonstrating apparent phase-based differences in muscle metabolic adaptation to SIT. Follicular phase-based training also showed significantly greater remodeling of extracellular matrix proteins (especially the major collagen types as for example; COL3A1, COL4A1-3, COL5A3, COL8A1), which was paralleled by the molecular chaperone SERPINH1 – required for collagen protein folding (Ito & Nagata, 2017) – as one of the most regulated proteins.

The suppression of mitochondrial pathways for luteal phase-based training was associated with a ≈3% impairment of V̇O_2max_, whereas it was not only maintained with follicular phase-based training but also paralleled by improved exercise capacity during the ramp test (within-subject CV of 2 and 3% for ramp exercise capacity and V̇O_2max_ in our lab). Although the decline in V̇O_2max_ for luteal phase-based training may appear small, this nevertheless is a meaningful change for highly-trained individuals (Joyner & Coyle, 2008). In other trained cohorts subjected to SIT, typically males, V̇O_2max_ generally remains unchanged (Hostrup & Bangsbo, 2017) as also observed in the follicular phase-based training group. While one could speculate that preceding training load influenced the performance measurements in our study, this is unlikely considering the recovery from last training until measurements as shown in other athlete cohorts (Davies et al., 1984; Yoon et al., 2007). SIT is well-tolerated even in untrained individuals (Little et al., 2010) and high-frequency training leading up to ramp testing do not influence V̇O_2max_ measurements in well-trained individuals (Billat et al., 1999). Our results are also coherent across the physiological outcomes and muscle proteome adaptations, which collectively suggest that follicular phase-based SIT is superior to luteal phase-based SIT under the settings in question.

Our findings highlight the menstrual cycle phase as an important physiologic confounder for muscle adaptations to exercise training and suggests phase-specific training as a potentially valuable strategy in optimizing training schedules for female elite athletes. This should not discourage female inclusion in sports science research. Rather, it emphasizes the imperative for further investigation. Implementing phase-based SIT would require careful tracking of each athlete’s menstrual cycle, with the understanding that certain individuals exhibit more pronounced within-subject differences in both hormone fluctuations and length of the menstrual cycle (Dam et al., 2022; McNulty et al., 2020; Mihm et al., 2011; Oosthuyse & Bosch, 2010). Nevertheless, given that pronounced muscle remodeling and phenotypic changes apparently occur after just one cycle of phase-based SIT this should encourage follow-up studies over several cycles where an even greater response could be expected.

## Materials and Methods

### Experimental design

In this study, we investigated the comprehensive protein response in skeletal muscle to phase-based high-frequency SIT during the menstrual cycle in female athletes. The study was a longitudinal, randomized parallel-group study. Participants performed two trial days (experimental day 1 and 2), before being randomized to a training intervention with either follicular phase-based training (FB) or luteal phase-based training (LB), lasting the length of one menstrual cycle for the individual participant (28 ± 2 days). 48-72 hours after completing the training intervention, participants completed two trial days (experimental day 3 and 4) identical to the trial days prior to the training intervention (Fig. **1**a). We hypothesized, that FB would be beneficial for both V̇O_2max_ and 30-min time trial performance. Secondarily, we anticipated greater adaptations in proteins involved in oxidative metabolism for FB compared to LB. All trials were conducted at the Department of Nutrition, Exercise and Sports, University of Copenhagen, Denmark between October 2020, and July 2022.

### Participants

The study was conducted in accordance with the standards set by the 2013 version of the Declaration of Helsinki and was approved by the regional ethics committee of Copenhagen, Denmark (H-20052639). All participants were informed about possible risks involved and gave their oral and written consent before conclusion. Inclusion criteria were healthy eumenorrheic female athletes not using hormonal contraceptives and with a regular menstrual cycle, age 18-45 years, V̇O_2max_ ≥50 mL·kg^-1^·min^-1^ (≤10% due to biological and technical variation) and ≥3 training sessions weekly. The included females performed primarily running, cycling, swimming and CrossFit. Exclusion criteria were chronic disease, injury on the musculoskeletal system, smoking, menstrual cycle >35 days, luteal phase-deficiency, anovulatory cycles, or pregnancy.

### Assessment of eligibility criteria

In the current study, we defined a regular menstrual cycle based on following criteria: a menstrual cycle between 21-35 days (Ritchie et al., 2015), positive ovulation, and serum progesterone concentrations above the lower threshold of 16 nmol·L^-1^ (Xiao et al., 2014). To verify this, and to track the transition between the follicular and luteal phase, participants completed calendar-based counting and urinary LH measurement. All females used calendar-based counting and reported the length of three full menstrual cycles prior to the intervention. In addition, previous users of hormonal contraceptives (four to six months prior) needed a positive ovulation test before baseline measurements to prevent anovulatory cycles. After baseline measurements all included participants tracked two full menstrual cycle by calendar-based counting and ovulation test. The onset of menses was registered as day 1, which indicated the change from luteal to follicular phase, as well as the day of positive ovulation test indicated the change from the follicular to luteal phase. The participants started ovulation testing from day 9 of the menstrual cycle until a positive test result occurred. The day of the positive ovulation test determined the scheduling of the subsequent trial period for the individual participant. A familiarization day was placed 1-5 days after positive ovulation test and both trial days pre and post the training intervention were placed 5-8 days after positive ovulation (mid-luteal phase). The trial days were placed in the mid-luteal phase to ensure that the performance tests were performed where the estrogen and progesterone concentrations are stable. To avoid including luteal deficient subjects in the project, participants with serum progesterone concentration of <16 nmol·L^-1^ in the mid-luteal phase (on trial days) were excluded for further data analysis (Yu et al., 2012). If all inclusion criteria and none of the exclusion criteria were met, participants were familiarized with the testing protocols on a separate day prior to the experimental days. At this familiarization day, a muscle biopsy was collected at rest from the vastus lateralis muscle using a Bergström biopsy needle through an incision made in the skin under local anesthesia (20 mg·mL-1 Xylocaine; AstraZeneca). Muscle biopsies were immediately rinsed in ice cold saline (9 mg/mL, Fresenius Kabi, Sweden) and dissected free of visible fat, blood, and connective tissue, before being frozen in liquid nitrogen and stored at −80°C for later analysis.

Furthermore, we assessed eligibility criteria by an incremental test to exhaustion on a bicycle ergometer (Lode Excalibur Sport, 10.10.1, The Netherlands, Groningen) for determination of V̇O_2max_ using an online breath-by-breath gas analyser system (Oxycon Pro, Viasys Healthcare, Germany, Hoechberg). The incremental test was preceded by a 12 min lead-in period of 4 min at 75, 100 and 125 W, respectively, followed by an increase in resistance by 25 W·min^-1^ until exhaustion. Incremental peak power output (iPPO) was defined as the highest workload (W) during the incremental test. After a recovery period of 10 min, participants had a 30-s lead-in period of 75 W, followed by an increase in workload to 110% of iPPO until exhaustion to maximize the likelihood of achieving true V̇O_2max_. Indirect calorimetry during the incremental test enabled measurement of substrate oxidation (carbohydrate and fat oxidation) at each of the three submaximal loads during the warm-up period (Peronnet & Massicotte, 1991). For the V̇O_2max_ measurements, the pre incremental test was conducted as the baseline measurement to ensure that the inclusion criteria for each participant were met, which was why the timing of this test not occurred in de exact same phase for all participants. However, it was not assumed that different hormone concentrations on the respective test days influenced the results of the incremental test, since previous studies on moderately trained and trained females have shown that V̇O_2max_ and iPPO were unchanged throughout the menstrual cycle (Casazza et al., 2002; Dean et al., 2003; Goldsmith & Glaister, 2020; Gordon et al., 2018; Jurkowski et al., 1981; Smekal et al., 2007).

### Outcome measurements and sample size

The main response outcome measure was performance in 30 min time trial and oxidative capacity. Other outcomes were V̇O_2max_, exercise capacity defined (iPPO), as well as muscle adaptations which were investigated by western blotting and MS-based proteomics. Sample size was determined for the main outcome measure in GPower 3.1.9.3 with an α-level of 005 and β-level of 0.8 for a linear mixed model repeated measures design which resulted in a required sample size of 12 participants per group. Participants were randomized with minimization for relative V̇O_2max._

### Training

The training intervention consisted of 8 SIT sessions on a spinning bike (Body Bike Indoor Cycle, W014). FB performed high-frequency SIT in the follicular phase (6 training sessions) and low frequency SIT in the luteal phase (2 training sessions). In contrast, LB performed low frequency SIT training in the follicular phase (2 training sessions) and high-frequency SIT in the luteal phase (6 training sessions) (Fig. **1**b). A training session consisted of a 5-min warm-up period at a self-selected resistance. Then, 6 × 30-s all out sprints were performed on a spinning bike followed by 3 min of rest. At the last training session, a biopsy was collected from the m. vastus lateralis at rest. Subsequently, a V̇O_2max_-ramp test to exhaustion was performed on a cycle ergometer, and participants completed a training session on a spinning bike consisting of 5 × 30-s all out bouts interspersed by 3 min of rest. In FB, participants completed 7 (n=2) or 8 (n=11) training sessions over 28±2 days, while in LB, participants completed 7 (n=3) or 8 (n=9) training sessions over 28±2 days. All missed or misplaced training sessions were attributed to menstrual cycle variations.

### Experimental protocol

On the experimental days, participants arrived at the lab after consuming a standardized meal from home. Body composition of was determined by dual-energy X-ray absorptiometry (DXA; Lunar iDXA, GE Healthcare, GE Medical systems, Belgium) and a blood sample was collected from the brachial vein in a 3.5 mL LH Lithium Heparin Sep tube to measure serum progesterone concentration. The blood sample was analyzed at Clinical Biochemistry Department 3011, Rigshospitalet, Copenhagen. Then, participants performed a standardized warm-up period on a bicycle ergometer (Lode Excalibur Sport, 10.10.1, The Netherlands, Groningen) consisting of 3 × 4 min at a resistance corresponding to 30, 50 and 70% of V̇O_2max_, respectively, followed by a recovery period of 5 min. After the warm-up period, participants completed a 30-min time trial on a bike ergometer. The protocol consisted of a 30-s lead-in period of 75 W to reach 80-90 RPM, followed by a 30-min isokinetic test with a fixed cadence of 90 RPM. During the time trials, participants aimed to perform all out while remaining seated. MPO was determined as the average power output over the whole 30-min time period.

### Experimental procedures

#### Proteomic sample preparation

Freeze-dried muscle tissue was powdered and then lysed with 4% sodium dodecyl sulfate (SDS), 100m M Tris at pH 8.5, using a BeatBox homogenizer (Preomics). The resulting lysate was immediately boiled for five minutes, cooled on ice, and subjected to tip-probe sonication (30:30s on/off, 15 cycles). Samples were centrifuged at 16,000 g for 10 minutes and the supernatant obtained was used for protein concentration determination employing a bicinchoninic acid (BCA) assay. For further processing, 50 µg of protein lysate was reduced with 10mM tris-(2-carboxyethyl) phosphine (TCEP) and alkylated with 40 mM 2-chloroacetamide (CAA), for 5 min at 40°C. An overnight on-bead digestion using a combination of trypsin (1:100 enzyme:protein) and lysC (1:500 enzyme:protein) was carried out via protein aggregation capture (PAC) on a KingFisher Flex robot (Thermo), as previously described (Batth et al., 2019). Protein digestion was quenched by adding 1% trifluoroacetic acid (TFA) and the peptides were desalted on C18 cartridges (Sep-Pak, Waters). Peptides were eluted with 50% acetonitrile (ACN), followed by vacuum drying to determine the concentration by measuring 280/260 absorbance in a NanoDrop spectrophotometer (Thermo Scientific). Finally, 200 ng of peptides were loaded onto Evotips previously equilibrated and ready for measurement.

#### Mass spectrometry analysis

The samples underwent separation using a 15-centimeter column with a diameter of 150 micrometers, packed with C18 beads (1.5 μm, Pepsep), on an Evosep ONE HPLC. The system utilized the default 30-SPD method, allowing for the processing of 30 samples per day, with the column temperature maintained at 50°C. After elution, peptides were injected into a timsTOF Pro 2 mass spectrometer (Bruker) using a CaptiveSpray source and a 20-micrometer emitter, operating in diaPASEF mode (Meier et al., 2018). For the global proteome profiling, the mass spectrometry data covered a range from 100 to 1700 m/z. During the collection of MS/MS data, each diaPASEF cycle lasted 1.8 seconds, covering an ion mobility range of 1.6-0.6 1/K0. The diaPASEF method employed a long-gradient approach, consisting of 16 diaPASEF scans with two 25 Da windows per ramp, setting a mass range of 400.0-1201.0 Da and a mobility range of 1.43-0.60 1/K0. Collision energy varied according to ion mobility, and decreased linearly from 59 eV at 1/K0 = 1.3 to 20 eV at 1/K0 = 0.85 Vs cm_2_. Calibration of ion mobility was performed using three Agilent ESI-L Tuning Mix ions (m/z 622.0289, 922.0097, and 1221.9906) with both accumulation and PASEF ramp times constant at 100 ms.

#### Mass spectrometry data processing

Raw MS files were analyzed in Spectronaut (version v.18.6, Biognosys) using a spectral library-free approach (directDIA+) with default settings against the reviewed human protein reference database; SwissProt human FASTA files (January 2023).

### Statistical Analysis

The statistical analysis was carried out in SPSS version 28 (IBM Software, IL, US). Normal distribution of data was confirmed by Q-Q plots and Shapiro Wilks test. A 2×2 linear mixed effects model with time (pre/post) and group (FB/LB) as fixed factors and participant as random factor was used to examine differences within and between groups over the intervention period. Data is presented as mean *±* standard deviation, and effect sizes as mean change with *±* 95% confidence interval. P-values are presented to represent probability.

### Bioinformatics

Data was filtered for 70% valid values in at least one group (FB or LB), resulting in 4155 proteins. Values were log_2_-transformed and medianscaled, followed by annotation with gene ontology terms (GO:BP, GO:CC, GO:MF) and keywords from the Uniprot database. Mitochondrial proteins were annotated with MitoCarta 3.0 (Rath et al., 2021). Data were analyzed in R (v.4.3.2, Foundation for Statistical Computing, Vienna, Austria) using the limma package (v. 3.52.4) (Ritchie et al., 2015). To control for multiple testing, a ∏-score was calculated based on statistical significance (p-value) and biological relevance (log_2_fold change) (Xiao et al., 2014). For gene set enrichment analysis, the R package clusterProfiler (Yu et al., 2012) (v. 4.4.4) was used and term p-values were corrected with the Benjamin-Hochberg procedure.

## Author contributions

Conception and design of the study: JK, KJJ, SJ, MH. Performed experiments: JK, KJJ, LBT.

Performed analyses: JK, SJ, JPQ, ASD, MH. Interpreted results: all.

Drafted the manuscript: JK, SJ, MH.

Edited and revised the manuscript: JK, SJ, JPQ, ASD, JB, MH. Read and approved final version: all.

## Data availability

The mass spectrometry proteomics data have been deposited to the ProteomeXchange Consortium via the PRIDE(Perez-Riverol et al., 2022) partner repository with the dataset identifier PXD051852.

## Additional information

Correspondence and requests for material should be addressed to MH.

